# Mpox vaccine and infection-driven human immune signatures

**DOI:** 10.1101/2023.03.07.23286701

**Authors:** Hallie Cohn, Nathaniel Bloom, Gianna Cai, Jordan Clark, Alison Tarke, Maria C Bermúdez-González, Deena Altman, Luz Amarilis Lugo, Francisco Pereira Lobo, Susanna Marquez, PVI study group, Jin-Qiu Chen, Wenlin Ren, Lili Qin, Shane Crotty, Florian Krammer, Alba Grifoni, Alessandro Sette, Viviana Simon, Camila H. Coelho

## Abstract

**Background:** Mpox (formerly known as monkeypox) outbreaks outside endemic areas peaked in July 2022, infecting > 85,000 people and raising concerns about our preparedness against this emerging viral pathogen. Licensed and approved for mpox, the JYNNEOS vaccine has fewer side effects than previous smallpox vaccines and demonstrated efficacy against mpox infection in humans. Comparing JYNNEOS vaccine- and mpox-induced immunity is imperative to evaluate JYNNEOS’ immunogenicity and inform vaccine administration and design.

**Methods:** We examined the polyclonal serum (ELISA) and single B cell (heavy chain gene and transcriptome data) antibody repertoires and T cells (AIM and ICS assays) induced by the JYNNEOS vaccine as well as mpox infection.

**Findings:** Gene-level plasmablast and antibody responses were negligible and JYNNEOS vaccinee sera displayed minimal binding to recombinant mpox proteins and native proteins from the 2022 outbreak strain. In contrast, recent mpox infection (within 20-102 days) induced robust serum antibody responses to A29L, A35R, A33R, B18R, and A30L, and to native mpox proteins, compared to vaccinees. JYNNEOS vaccine recipients presented comparable CD4 and CD8 T cell responses against orthopox peptides to those observed after mpox infection.

**Interpretation:** JYNNEOS immunization does not elicit a robust B cell response, and its immunogenicity may be mediated by T cells.

**Funding:** Research reported in this publication was supported, in part, by the National Cancer Institute of the National Institutes of Health under Award Number U54CA267776, U19AI168631(VS), as well as institutional funds from the Icahn School of Medicine.

## Introduction

Mpox (formerly known as monkeypox) is a zoonotic infection caused by an orthopoxvirus (OPXV). This viral genus includes smallpox, which has dramatically affected humanity for centuries. As we progressed toward its eradication in the 1980s, mpox became the most concerning OPXV for humans, causing multiple outbreaks in Central and West Africa since 1970, and its first outbreak outside Africa (in the US) in 2003^1^. The 2022 global mpox outbreak infected > 85,000 people, providing an unprecedented opportunity to study human immune responses to mpox outside Africa. While the World Health Organization reported that disease incidence has decreased by 90% compared to the outbreak’s peak in July 2022^2^, mpox transmission continues in Africa and Latin America^3^.

The recently implemented US mpox vaccination strategy made two smallpox vaccines available to healthcare workers and individuals 18 years of age and older: ACAM2000 (a second-generation live-replicating vaccinia strain; Emergent Product Development, Gaithersburg, MD, USA) and JYNNEOS (a live non-replicating modified Ankara strain of vaccinia; Bavarian Nordic, Hellerup, Denmark), which is the only vaccine approved for mpox prevention. Licensed in the US in 2019, JYNNEOS is a third-generation smallpox vaccine designed to elicit fewer side effects than ACAM2000^4^.

To date, robust data demonstrating JYNNEOS protection has only been shown in animal models^5^. Initial clinical trial data from the UK^6^ and the US^7^ demonstrated that JYNNEOS efficacy in humans could reach 78% and 87%, respectively, although these studies did not assess how the efficacy data could be impacted by behavioral differences between the vaccinated and unvaccinated individuals. However, while the immunogenicity of the ACAM2000 vaccine has been investigated^8,9^, little is known about how JYNNEOS induces immune protection in humans^10,11^.

Human smallpox vaccination responses are highly mediated by antibodies^12–16^ that can remain elevated for years^15,17–22^. However, despite this prolonged smallpox protection, the high frequency of conserved antigens shared between smallpox and mpox^13,14,23^, and the fact that vaccinia-induced antibodies protect non-human primates from mpox infection^24^, vaccinia (smallpox) vaccination does not provide complete protection of mpox symtpons in humans^14,25^. This suggests that mpox immunity might be driven by mpox-specific antigens or that protection is not strongly antibody-mediated. Finally, it is unclear how JYNNEOS shapes the human antibody repertoire or how efficiently the antibodies of vaccinated or convalescent participants neutralize the current mpox outbreak strains.

Given the prolonged antibody responses of older smallpox vaccines and high sequence homology among common OPXVs^4,26^, JYNNEOS was expected to elicit neutralizing immune responses against the 2022 outbreak strain. However, initial data suggest low humoral immunogenicity and low virus neutralization potential^11^. Furthermore, no information is available on how the vaccine alters the antibody repertoire at the immunoglobulin gene level to enable rapid production of antibody-producing B cells targeting mpox epitopes upon future encounters. Thus, the characterization of JYNNEOS-induced antibodies is imperative.

Here, we have examined the human antibody repertoire induced by JYNNEOS in US adult vaccinees at the single B cell and serum levels. This is the first report of a single-cell human antibody repertoire induced by mpox vaccination and the first systematic examination of T cell activation after JYNNEOS vaccination. Importantly, we demonstrate that JYNNEOS immunity may be primarily T cell-mediated, with minimal activation of humoral immunity.

## Methods

### Participant demographic and sample collection

The study protocols for clinical specimen collection from convalescent and post-vaccination individuals by the Personalized Virology Initiative were reviewed and approved by the Mount Sinai Hospital Institutional Review Board (IRB-16-16772, IRB-16-00791). All participants provided written informed consent before specimens and clinical information were collected. Permissions to store and share biospecimen were also obtained from all participants. All specimens were coded before processing and analysis. Whole blood was collected through venipuncture into serum separator tubes and ethylenediaminetetraacetic acid tubes. Serum and plasma were stored at −80 °C until analysis. Peripheral blood mononuclear cell (PBMC) isolation was performed by density gradient centrifugation using SepMate tubes (STEMCELL Technologies, Cambridge, MA, USA). Samples were cryo-preserved in liquid nitrogen until analysis.

Samples were collected from 16 participants. Ten were vaccinated with JYNNEOS and six were diagnosed with mpox infection **(Table 1)**. Blood samples were collected from seven vaccinees before vaccination to serve as a baseline. Vaccinee samples were collected on average 35 days post-dose one (range: 6–60 days) and 13 days post-dose two (range: 7– 40 days). Mpox convalescent samples were collected at an average of 55 days post-infection (range: 20–102 days; **(Table 1)**.

**Table 1.** Subject demographics and immunization routes of JYNNEOS vaccine recipients and mpox-convalescent participants.

| Subject ID | Sex | Age | Race | Orthopoxviral exposure | IR-1<sup>st</sup> Dose | IR-2<sup>nd</sup> Dose |
| --- | --- | --- | --- | --- | --- | --- |
| **V1** | M | 21-30 | C | JYNNEOS | Subcutaneous | Intradermal |
| **V2** | M | 31-40 | C | JYNNEOS | Subcutaneous | Intradermal |
| **V3** | M | 41-50 | C | JYNNEOS | Subcutaneous | - |
| **V4** | M | 41-50 | H | JYNNEOS | Subcutaneous | Subcutaneous |
| **V5** | M | 31-40 | H | JYNNEOS | Subcutaneous | Intradermal |
| **V6** | M | 41-50 | C | JYNNEOS | Subcutaneous | Intradermal |
| **V7** | M | 31-40 | C | JYNNEOS | Subcutaneous | Intradermal |
| **V8** | M | 41-50 | C | JYNNEOS | Subcutaneous | Intradermal |
| **V9** | M | 41-50 | C | JYNNEOS | Intradermal | - |
| **V10** | M | 31-40 | C | JYNNEOS | Subcutaneous | Subcutaneous |
| **C1** | M | 41-50 | AA | Mpox Infection | - | - |
| **C2** | F | 31-40 | AA | Mpox Infection | - | - |
| **C3** | M | 31-40 | C | Mpox Infection | - | - |
| **C4** | M | 41-50 | C | Mpox Infection | - | - |
| **C5** | M | 31-40 | C | Mpox Infection | - | - |
| **C6** | M | 51-60 | E | Mpox Infection | - | - |

IR= immunization route. C = Caucasian, H = Hispanic, AA= African American E=Ecuadorian.

**Table 2.**
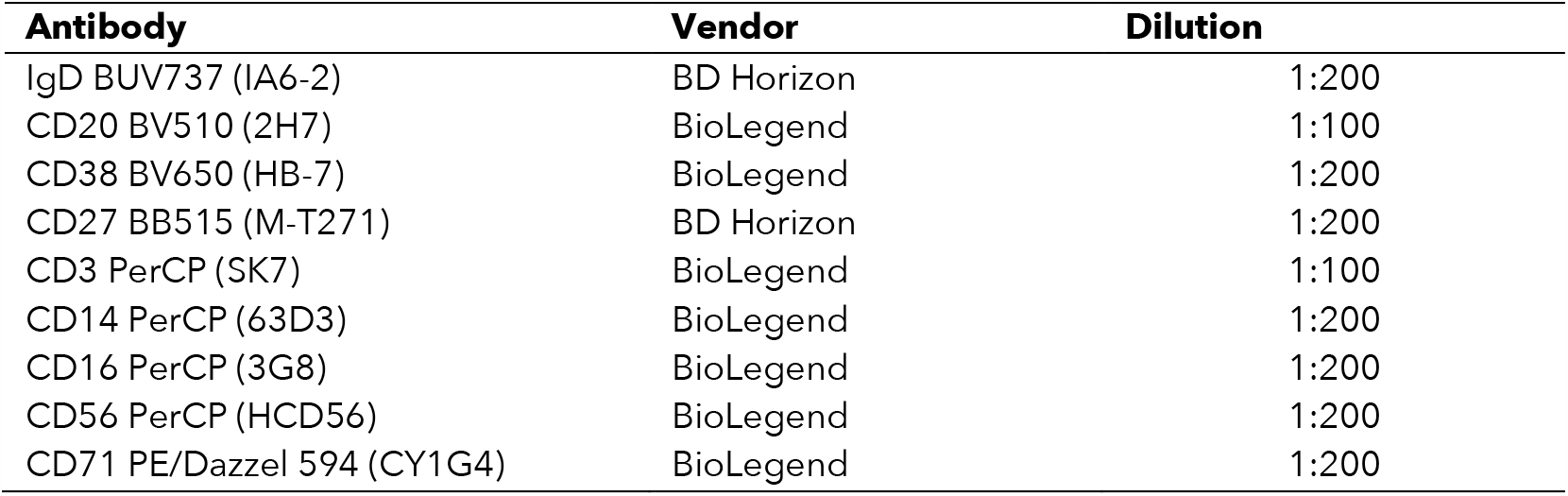
Antibodies used for flow cytometry and cell sorting.

| Antibody | Vendor | Dilution |
| --- | --- | --- |
| IgD BUV737 (IA6-2) | BD Horizon | 1:200 |
| CD20 BV510 (2H7) | BioLegend | 1:100 |
| CD38 BV650 (HB-7) | BioLegend | 1:200 |
| CD27 BB515 (M-T271) | BD Horizon | 1:200 |
| CD3 PerCP (SK7) | BioLegend | 1:100 |
| CD14 PerCP (63D3) | BioLegend | 1:200 |
| CD16 PerCP (3G8) | BioLegend | 1:200 |
| CD56 PerCP (HCD56) | BioLegend | 1:200 |
| CD71 PE/Dazzel 594 (CY1G4) | BioLegend | 1:200 |

### Flow cytometry and cell sorting

Cryopreserved PBMCs from JYNNEOS recipients were thawed in Roswell Park Memorial Institute RPMI 1640 Medium w/ L-glutamine and 25 mM HEPES (Corning, Corning, NY, USA, 10-041-CV) supplemented with 10% fetal bovine serum (FBS) and 500 U of Benzonase Nuclease HC (MilliporeSigma, Burlington, MA, USA, 71206-25KUN) at 37°C. Cells were washed with cold FACS buffer (phosphate-buffered saline (PBS; Thermo Fisher Scientific, Waltham, MA, USA, 10010023) with 2% FBS), resuspended for counting, then aliquoted into 96-well plates (Corning 96-well Clear Polystyrene Microplates, Corning, #3788) and stained for plasmablasts defined as (Dump-(CD3, CD14, CD16, CD56), CD19+ IgD-, CD20-CD38hi, and CD71+) (**Figure S1**) in BD Horizon Brilliant Stain Buffer (BD Biosciences, San Jose, CA, USA, 566349) for 30 min at 4°C. Cells were then washed with cold FACS buffer and dead cells were detected using the LIVE/DEAD Fixable Blue Dead Cell Stain Kit (Thermo Fisher Scientific, L34962) in Dulbecco’s PBS with calcium and magnesium (Thermo Fisher Scientific, 14040133) at 4*°*C for 15 min. Cells were washed and resuspended in FACS buffer, then sorted into Eppendorf™ DNA LoBind Microcentrifuge Tubes (Thermo Fisher Scientific, 022431021) containing 12 μL of PBS with 0.05% bovine serum albumin (BSA) and directly loaded into a 10x Genomics Chromium Next GEM Chip K (10x Genomics, Pleasanton, CA, USA, PN-2000182) for analysis.

### Single-cell RNA sequencing library preparation

Sorted plasmablasts were processed using 10x Genomics kits (Next GEM Single Cell 5′ GEM Kit v2 Dual Index (PN-1000244), Chromium Next GEM Chip K Single Cell Kit (PN-1000286), Chromium Single Cell Human BCR Amplification Kit (PN-1000253), Library Construction kit (PN-1000190), and Dual Index Kit TT Set A (PN-1000215)) according to the manufacturer’s protocols. We used cDNA to prepare B cell receptor (BCR) and 5′ gene expression libraries. All quantifications were performed on a TapeStation 2200 (Agilent, Santa Clara, CA, USA). Libraries were sequenced on a NovaSeq 6000 at sequencing read depths of 5,000 and 50,000 for VDJ and gene expression libraries, respectively.

### Bioinformatic analyses of BCR sequencing

Raw sequencing reads were processed with Cell Ranger 7.0.1 (10x Genomics) and reference VDJ germline sequences were retrieved from IMGT (2022-12-28). The assembled contigs were analyzed with the nextflow pipeline nf-core/airflow (https://nf-co.re/airrflow commit 072a40bf6d8352f9dc33456d69b9156ffb2906bd), using an Immcantation (http://www.immcantation.org) docker container (build 515029fbe323e7479190f14ebcf188ec76f6d90cfea7ce3a89d17f24ee7ded35, available at DockerHub) and the same reference germlines used with Cell Ranger. The pipeline performs the following processing steps: gene assignment with IgBLAST (version 1.20.0), quality filtering (retaining sequences with concordant loci in the v_call and locus fields identified by IgBLAST, with a sequence_alignment with a maximum of 10% Ns and a length of at least 200 informative nucleotides (not -, ., or N)), removal of non-productive sequences, removal of sequences with junction lengths that are not multiples of 3, single cell-specific quality filters (keeping cells with one heavy chain, removing sequences with the same cell_id, and sequence in different samples), and clustering of clonally related sequences. To identify clones, we used the hierarchical clustering method implemented in SCOper^27^ with a nucleotide Hamming distance threshold of 0.17. Differences in gene and isotype usage and mutation frequency were determined by *t*-test.

### Bioinformatic analyses of gene expression data

We used the Cell Ranger^28^ pipeline and the Seurat R^29^ package for single-cell transcriptome analysis. The steps were as follows: fastq generation and demultiplexing, quality control at the library and individual cell levels, transcript quantification and normalization, dimensionality reduction, cell clustering, differential gene expression analysis, and data visualization. We ran the Cell Ranger pipeline to obtain gene expression count matrices from the raw sequencing data using standard parameters and the expected number of cells for each sample set as the number of sorted cells (using the “--expect-cells” parameter). The gene expression data and metadata for each cell were used to create Seurat objects. We removed cell barcodes with < 200 or > 2,500 genes detected (nFeature_RNA), as well as those with > 10% mitochondrial content. The number of cell barcodes was then downsampled to match the sample with the least barcodes. We normalized each subsample using regularized negative binomial regression as provided by the SCTransform function, resulting in a list of normalized Seurat objects. The top 3,000 most variable genes across each comparison were selected using the SelectIntegrationFeatures function and used for sample integration using the PrepSCTIntegration, FindIntegrationAnchors, and IntegrateData functions. We reduced the dimensionality of the data using the uniform manifold approximation and projection (UMAP) technique on the 30 largest principal components computed by principal component analysis with the RunUMAP function and used this data structure to find neighbours and cluster cell groups using the FindNeighbors and FindCluster functions, respectively. We computed the differentially expressed genes in each group using the Wilcoxon rank-sum test, followed by Benjamini-Hochberg false discovery rate (FDR) correction. Genes with FDR < 0.01 and log fold change (FC) > 0.2 were considered differentially expressed in an experimental group. We observed a bimodal distribution in the total reads mapping to genes encoding ribosomal proteins, and a considerable fraction of these genes were differentially expressed. For this reason, we repeated the pipeline described above with the 103 ribosomal genes removed from the count matrices before sample normalization.

### Recombinant proteins

To express proteins derived from the Zaire-96-I-16 strain of mpox, cDNAs encoding residues 1–275 of E8L (GenBank: 72551515), 22–146 of A30L (GenBank: 72551549), 1–110 of A29L (GenBank: 72551548), and 58–181 of A35R (GenBank: 72551555) were synthesized along with a GGGSGGGS linker and a 10×His tag and cloned into the mammalian expression vector pcDNA3.4. These expression vectors were transiently transfected into HEK293F cells with polyethylenimine. Proteins were purified from filtered cell supernatants with Ni-NTA resin before additional purification by size-exclusion chromatography using a Superdex 200 Increase 10/300 GX column (GE Healthcare, Boston, MA, USA) in PBS (pH 7.4) containing 0.005% Tween 80 and 10% trehalose. These recombinant mpox proteins are available for purchase from ACROBiosystems (Newark, DE, USA; E8L, E8L-M52H3; A30L, A3L-M5243; A29L, A2L-M52H3; A35R, A3R-M52H3).

Recombinant vaccinia virus (VACV) proteins B5 (40900-V08H), L1R (40903-V07H), B18R/B19R (40020-V08B), A27L (40891-V07E), and A33R (40896-V07E), from the Copenhagen strain of VACV, were purchased from Sino Biological (Beijing, China).

### Enzyme-linked immunosorbent assays (ELISAs)

Serum antibody titers were quantified via ELISA using the recombinant proteins listed above and lysates derived from cells infected with the current US outbreak strain of mpox. In addition, lysates of mpox infected HRT-18 cells were also used to probe polyclonal antibody responses. Following lysis in 1% SDS solution and freezing at –80C, the SDS was removed using the SDS-out SDS Precipitation Kit (Thermo Fisher Scientific, 20308) according to the manufacturer’s instructions. ELISAs were performed in MultiScreen® 96-well ELISA high binding plates (Millipore, MSEHNFX40). Plates were coated with 50 μL of 2 μg/mL recombinant protein, except for A30L and E8L, which were coated at 1 μg/mL. Inactivated SDS-free mpox cell lysates were coated at 5 μg/mL. All plates were coated for 1 hour in a 37°C incubator, then blocked for 1 hour at room temperature in PBS supplemented with 0.1% Tween 20 (PBST; Thermo Fisher Scientific ref. J61419.K3) and 3% milk powder (AmericanBio, Canton, MA, USA, AB1010901000). Heat-inactivated serum samples were diluted 1:40 in PBST containing 1% milk powder and then serially diluted 1:3. The blocking solution was removed from the plates and the serum dilutions were added. After incubation for 2 hours at room temperature, plates were washed three times with PBST using a BioTek 405 TS microplate washer (Agilent). After washing, 100 μL of Anti-Human IgG (Fab specific)−Peroxidase secondary antibody (produced in goat; Sigma-Aldrich, St. Louis, MO, USA, A0293, RRID: AB_257875) diluted 1:3,000 in PBST with 1% milk powder was added. After a 1-hour incubation at room temperature, the plates were washed three times with PBST and 100 μL SIGMA*FAST*^*TM*^ OPD (Sigma-Aldrich, P9187) was added. After a 10-minute incubation at room temperature in the dark, the reaction was stopped by adding 50 μL of 3 M hydrochloric acid (Thermo Fisher Scientific, S25856) to each well. The optical density at 490 nm of each plate was measured using a BioTek Synergy H1 Multi-Mode Reader (Agilent). The area under the curve (AUC) was then calculated for each plate and plotted using GraphPad Prism 9 software (GraphPad Software, San Diego, CA, USA).

### Combined activation-induced marker (AIM)/intracellular cytokine staining (ICS) assays to detect antigen-specific T cell responses

We investigated orthopox-specific T cell responses using previously described peptide pools^30^. The OPXV peptides were based on experimentally defined CD4 and CD8 epitopes from IEDB (www.IEDB.org). Peptides were synthesized as crude materials (TC Peptide Lab, San Diego, CA, USA), pooled into OPXV CD4 and OPXV CD8 mega pools, and sequentially lyophilized^30^.

To assess OPXV-specific T cell responses, PBMCs were cultured with the OPXV-specific peptide mega pools (1 μg/mL). An equimolar amount of dimethyl sulfoxide (DMSO) was added to duplicate wells as a negative control, and phytohemagglutinin-L (1 μg/mL, Millipore, Saint Louis, MO, USA 431784) was used as a positive control. Stimulated cells were incubated with CD40 (1:133, Miltenyi Biotec, Gaithersburg, MD, USA, 130-094-133) and CXCR5 BV650 (1:100, BD Biosciences, 740528) antibodies at 37°C in 5% CO_2_ for 26 hours. During the last 4 hours of incubation, a combination of Golgi-Plug containing brefeldin A, Golgi-Stop containing monensin (1:1000, both BD Biosciences), and CD137 APC antibodies (1:100, BioLegend, San Diego, CA, USA, 309810) was added. Membrane surface staining was performed for 30 minutes at 4°C with eBioscience™ Fixable Viability Dye eFluor™ 506 (1:1,000, Thermo Fisher Scientific, 65-0866-14) and the following antibodies: CD3 BUV805 (1:50, BD Biosciences, 612895), CD8 BUV496 (1:50, BD Biosciences, 612942), CD4 BUV395 (2:100, BD Biosciences, 564724), CD14 V500 (1:50, BD Biosciences, 561391), CD19 V500 (1:50, BD Biosciences, 561121), CD69 BV605 (4:100, BioLegend, 310938), CD137 APC (1:50, BioLegend, 309810), OX40 PE-Cy7 (1:50, BioLegend, 350012), CXCR5 BV650 (2:100, BD Biosciences, 740528), and CD154 (CD40 Ligand) Monoclonal Antibody (24-31), APC-eFluor™ 780, eBioscience (5:100, Thermo Fisher Scientific, 47-1548-42). Cells were then fixed with 4% paraformaldehyde (Sigma-Aldrich), permeabilized with saponin buffer (0.5% saponin (Sigma-Aldrich), 1% BSA, and 0.1% sodium azide), and blocked for 15 minutes with 10% human serum (Gemini Bio-Products, Sacramento, CA, USA) in saponin buffer. Intracellular staining was performed for 30 minutes at room temperature with the following antibodies: TNFα-PE (1:500, Thermo Fisher Scientific, 12-7349-82), IFNγ FITC (1:50, Thermo Fisher Scientific, 11-7319-82), IL4 BV711 (1:50, BD Biosciences, 564112), IL10 PE-Dazzle594 (1:100, BioLegend, 506812), and granzyme B Alexa 700 (1:100, BD Biosciences, 560213). Samples were run on a ZE5 Cell Analyzer (Bio-Rad, Hercules, CA, USA) and were analyzed with FlowJo 10.8.1 (Tree Star Inc., Ashland, OR, USA). Limit of detection (twice the upper 95% confidence interval of the geometric mean) and limit of sensitivity (LOS, twice the standard deviation from the median) calculations were based on the DMSO-only conditions for AIM and ICS. Responses were considered positive with a stimulation index > 2 and AIM LOS values of 0.02 and 0.04 for CD4 and CD8, respectively. For ICS, a stimulation index > 2 was considered positive when combined with LOS values of 0.01 for both CD4 and CD8.

## Results

B cell activation by immunization leads to their differentiation into short-lived, antibody-secreting plasmablasts. Single-cell sequencing of plasmablasts obtained 6–9 days after JYNNEOS vaccination demonstrated that neither one nor two doses significantly increased the number of variable heavy chain (VH) sequence clonotypes **(Figure S2A, Table S1)** or their diversity, as defined by three criteria **(Figure 1A)**. Two doses did not significantly alter the frequency of VH mutations **(Figure 1B)**, suggesting that at least shortly after each immunization, JYNNEOS vaccination does not sufficiently engage antigen-mediated germinal center responses to promote somatic hypermutation. Similarly, the length of complementarity-determining region 3 (CDR3) in the antibody heavy chain, an indicator of antigen binding specificity after vaccination^31,32^, was unaltered after JYNNEOS immunization **(Figure 1C)**. In pre-vaccinated (unvaccinated, and uninfected) participants, the antibody repertoire of naïve B cells mainly comprised IgM sequences **(Figure 1D)**, as recently demonstrated by Lanzavecchia and his group^33^. Compared to pre-vaccination, IgA levels were increased after one dose, which was accompanied by decreased IgM levels **(Figure 1D)**. Two doses of JYNNEOS did not alter immunoglobulin isotype frequencies compared to pre-vaccination **(Figure 1D)**. Similarly, one or two doses of JYNNEOS did not affect heavy chain mutation frequencies in the IgG, IgA, and IgM isotypes **(Figure 1E, Figure S2B)**. Compared to pre-vaccination, one or two doses of JYNNEOS did not significantly alter immunoglobulin heavy chain variable (IGHV) gene families **(Figure S2C)**. Although usage of *IGHV1-2, IGHV1-46, GHV4-39*, and *IGHV4-59* seemed slightly increased in two participants after the second dose **(Figure 2)**, these increases were not significant. Since no significant changes in plasmablast antibody gene expression were observed after one or two doses, we next expanded our search to characterize global gene expression patterns. After quality control **(Figure S4 and S5)**, we saw that in pre-vaccination CD19+ B cells, we identified transcripts commonly expressed in B cells, such as BANK1, BACH2, and ARHGAP24^34–36^ **(Figure S6)**. Given the high levels of ribosomal proteins expressed in total B cells, we also analyzed the dataset after removing these reads; however, the gene expression profiles identified differentially expressed B cell-related genes remained comparable **(Figure S7)**. Compared to their baselines, plasmablasts obtained from paired subjects post-dose one expressed high levels of CD74 **(Figure S6)**, which is expressed in antigen-experienced cells^37^ and was part of a gene cluster (cluster 1) that expanded after vaccination **(Figure S6)**. This change was expected since we analyzed plasmablasts, which are more differentiated than the total B cells taken pre-vaccination. However, and more importantly, the most differentially expressed genes after dose one and dose two were unchanged **(Figure S6)**, suggesting that a second dose of JYNNEOS did not significantly alter plasmablast gene expression.

**Figure 1.**
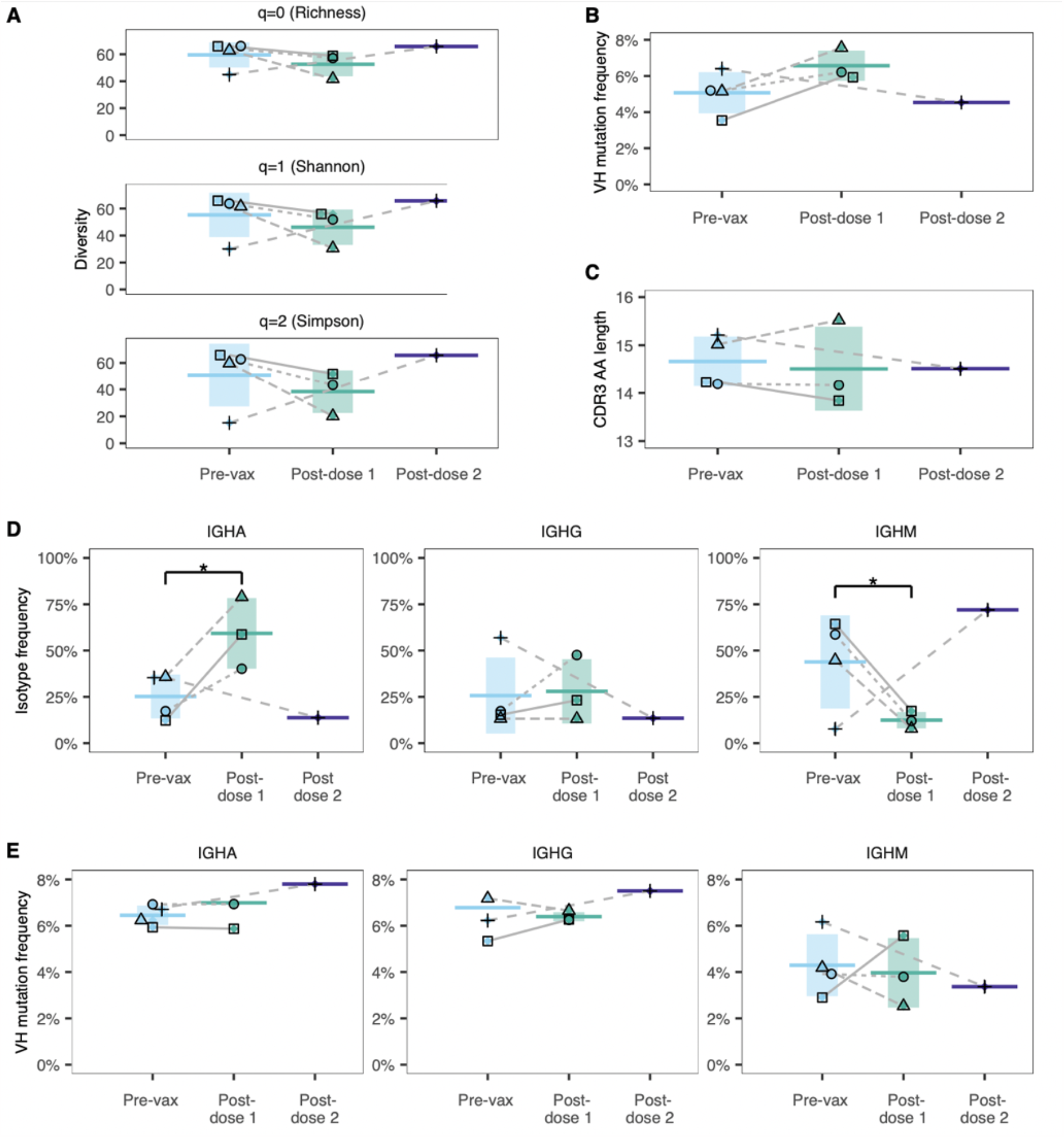
Antibody repertoires of single B cells after one and two doses of JYNNEOS. **(A)** Heavy chain variable (VH) sequence diversity was measured using three different parameters. Plasmablasts were collected 6–9 days after one or two doses of JYNNEOS. Total B cells (CD19+) from the same participants were collected before immunization and sorted for single-cell sequencing. **(B)** Frequency of VH mutations. **(C)** CDR3 amino acid (AA) lengths. **(D)** Frequencies of immunoglobulin isotypes. **(E)** Frequencies of heavy chain gene mutations according to immunoglobulin isotype. Statistical significance was evaluated with *t*-tests. Participants are identified by unique symbols.

**Figure 2.**
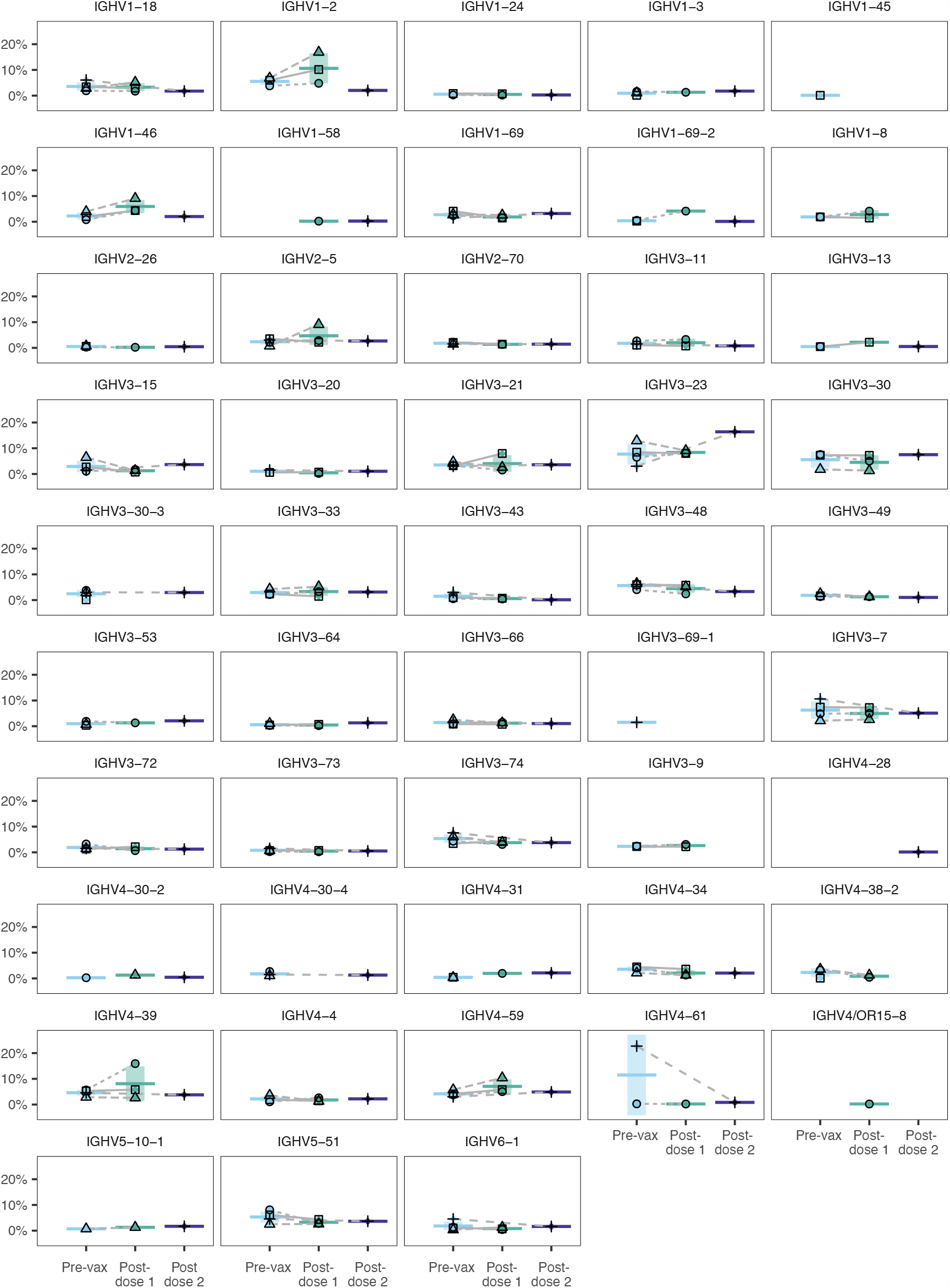
Variable gene usage in pre-vaccine B cells and plasmablasts after one or two doses of JYNNEOS. IGHV genes of the *IGHV-1*–*6* families are shown. Statistical significance was evaluated by *t*-test

As plasmablast responses seemed unaffected at the single-cell level, we also analyzed the antibodies they secrete into circulation. Since plasmablasts are short-lived, early responders that peak 6–9 days post-vaccine, we were also curious to evaluate vaccine responses up to 2 months post-immunization. We quantified serum IgG responses after immunization or infection. Initially, we expressed the mpox proteins A29L, A35R, A30L and E8L (strain Zaire-96-I-16), all known antibody targets^38^ that can induce B cell immunogenicity^39–44^. IgG responses to A29L and A35R proteins were increased only after infection **(Figure 3A–D)** and were undetectable or present at very low levels in the serum of vaccinees shortly after or up to 60 days after the first or second doses **(Figure S8)**. A30L was the only protein we analyzed that demonstrated JYNNEOS induced significant IgG response after two doses, with IgG levels comparable to those observed during convalescence. Responses to the E8L protein were detected in sera collected 18–60 days after one or two doses but not shortly after immunization **(Figure S8A–H)**.

**Figure 3.**
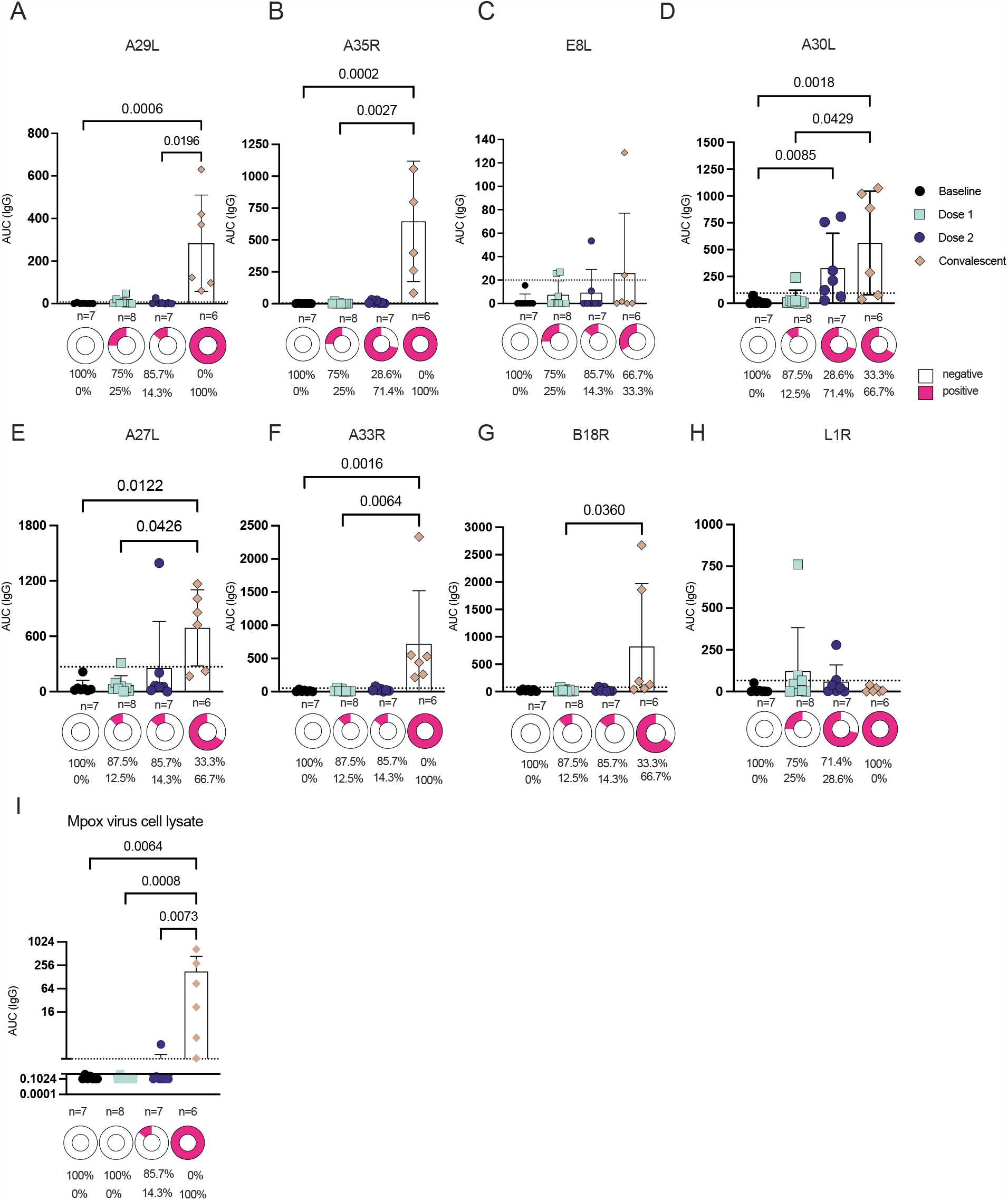
Serum antibody responses to recombinant mpox proteins and lysates from mpox-infected cells (A–D) Serum IgG responses to recombinant mpox proteins A29L (A), A35R (B), E8L (C), and A30L (D) at baseline and 18–60 days post-immunizations or 45–102 days post-infection. AUC, area under the curve. The donut graphs illustrate the proportion of samples in each group that were reactive to the recombinant proteins. Positivity was defined based on limit of detection (mean+ 3x SD of negative controls) **(E–H) Serum IgG responses to recombinant vaccinia proteins A27L (E), L1R (F), B18R (G) and A33R (H) at baseline and post-immunizations or -infection**. The day ranges are the same as A-D. **(I)** Serum IgG responses of vaccinated and convalescent participants to a lysate from mpox-infected cells. Dashed lines represent the limit of detection (defined as the mean + three standard deviations). Comparisons were performed using the Kruskal-Wallis test followed by Dunn’s multiple comparisons test (*p < 0.05, **p <0.01, ***p < 0.001). Supplementary Figure 8 shows additional IgG data from an earlier time point post-immunization.

As the high conservation of OPXVs results in the production of cross-reactive antibodies upon vaccination^13^, we next sought to assess serum antibody responses to vaccinia proteins previously identified as targets of human neutralizing antibodies^38^. One or two doses of JYNNEOS did not elicit robust IgG responses to the four proteins we tested (A27L, L1R, B18R and A33R). Infection, however, generated robust serum IgG responses against A33R and A27L **(Figure 3E–F)**. Only three convalescent individuals and none of the vaccinees had IgGs targeting B18R **(Figure 3G)**. Mpox infection did not generate antibodies against L1R, and only one subject was responsive to it after vaccination **(Figure 3H)**. As most convalescent subjects in this study also had human immunodeficiency virus (HIV), and HIV+ individuals are commonly (average of 18%)^45^ infected with the OPXV Molluscum contagiosum^46,47^, we sought to confirm whether their convalescent responses were mpox-specific or cross-reactive. We therefore tested the sera of HIV+ and HIV+/mpox+ subjects with the mpox protein A29L, which was very immunogenic in convalescents. Anti-A29L levels in co-infected subjects were found to be above the limit of detection compared with those with HIV only **(Figure S8I)**. This indicates that the responses raised were due to mpox infection alone.

Finally, we isolated mpox virus from an infected lesion derived from an infected individual during the 2022 outbreak and developed an ELISA to assess IgG responses to lysates from mpox-infected cells. The serum of vaccinees receiving one or two doses of JYNNEOS did not bind the mpox proteins in the lysate, while 4/6 convalescent participants elicited strong antibody responses **(Figure 3I)**.

Taken together, our results demonstrate that—unlike previous generations of smallpox vaccines—JYNNEOS does not induce a strong humoral response to OPXVs in humans. Human clinical trials indicate protection against mpox^6,7,48,49^. Therefore, we next investigated how JYNNEOS vaccination induces T cell immunity in humans compared to mpox infection. We measured the antigen-specific CD4+ and CD8+ T cell responses to JYNNEOS immunization or mpox infection using a previously reported^30^ pool of VACV peptides **(Figure 4)**. T cell responses were measured using a combined AIM/ICS assay (see **Figure S9A** for the gating strategy). CD4+ T cells showed pre-existing immunity before vaccination, with a higher frequency of positivity and increased reactivity after two doses and a response comparable to post-mpox infected samples (**Figure 4A–B)**. Circulating T follicular helper (cTFH) cell levels were significantly increased post-vaccination but not after mpox infection (**Figure 4C)**. CD8+ T cells displayed little to no evidence of pre-existing immunity, but robustly increased in response to two doses of JYNNEOS compared to baseline and after mpox infection (*p* < 0.05). Although they lacked pre-immunity, CD8+ T cell responses presented similar FC increases to CD4+ T cells in response to vaccination (**Figure 4D–E)**. The T cell responses involved the production of multiple cytokines, revealing a trend toward increased polyfunctionality post-vaccination or -infection (**Figure 4F**), including the presence of mixed T helper type (Th)1/(Th)2 cell phenotypes (**Figure S10**). Finally, no significant correlations were observed between the T cell and antibody responses (**Figure S11**) and the lower cTFH responses observed in convalescent samples were not correlated with the differences in the time following exposure between infected and vaccinated participants (**Figure S11**). Antibody responses to vaccination were also not associated with the day post-immunization, suggesting that although these samples were collected closer to exposure than the convalescent samples (**Figure S11**), this difference is likely not responsible for the superior antibody responses mounted against infection.

**Figure 4.**
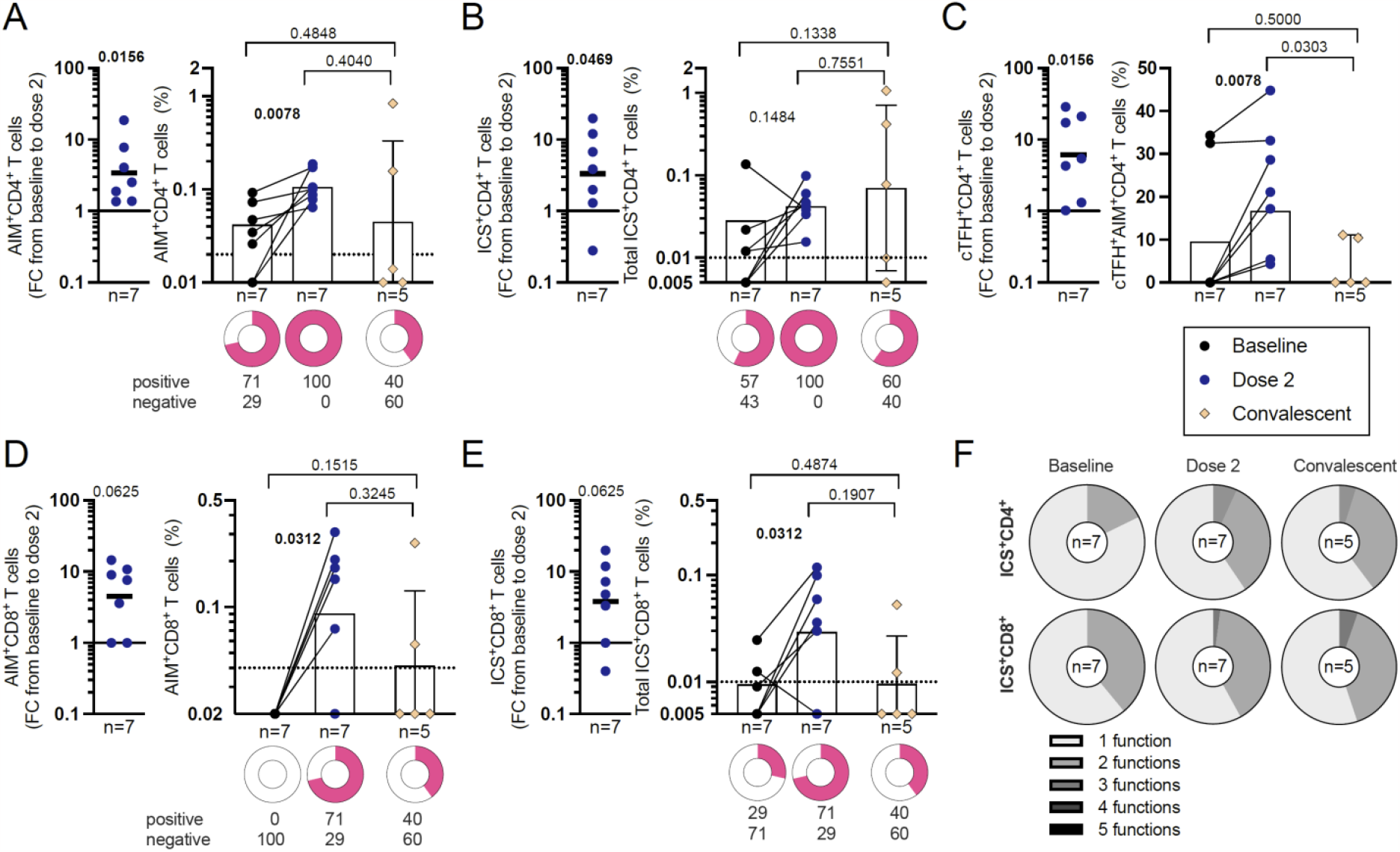
Human CD4+ and CD8+ T cell responses to JYNNEOS immunization or mpox infection. **(A–B)** CD4 T cell responses to the OPXV peptide pool 9–114 days after vaccination and the magnitudes of their AIM (A) or ICS (B) responses pre- and post-vaccination or 20–130 days post-mpox infection. FC, fold change. **(C)** OPXV-specific CD4 cTFH cell responses to the OPXV peptide pool after vaccination and AIM magnitudes pre- and post-vaccination or post-mpox infection. **(D–E)** CD8 T cell responses to the OPXV peptide pool after vaccination and the magnitudes of their AIM (D) or ICS (E) responses pre- and post-vaccination or post-mpox infection. **(F)** Functionality of OPXV-specific CD40L+ CD4 and CD69+ CD8 cytokine responses pre- and post-vaccination or post-mpox infection. Dashed lines represent the LOS. The significance of the FC was assessed by Wilcoxon signed rank test compared to a hypothetical median of 1. Baseline and post-dose two values were compared by one-tailed Wilcoxon test, baseline and convalescent samples were compared by one-tailed Mann-Whitney test, and dose two and convalescent were compared by two-tailed Mann-Whitney test. *P*-values are listed above the data points. The donut graphs represents the frequency of positivity based on the limit of detection. Supplementary Figure 9 shows the gating strategy for the T cell assay and the individual cytokine responses.

## Discussion

Human OPXV vaccines have an extraordinary history of successful protection from illness. The first-generation Dryvax vaccine prevented smallpox but caused severe side effects, leading to the development of second- (ACAM2000) and third- (JYNNEOS) generation vaccines. JYNNEOS is safe, immunogenic, and protective against smallpox in humans^50–52^. Consequently, it was authorized by the US Food and Drug Administration (FDA) for mpox prevention during the 2022 global outbreak. However, the immunogenicity and efficacy of third-generation vaccines designed to prevent smallpox have not been thoroughly evaluated, including their functional activities and, more importantly, their mechanism(s) of protection against mpox. To gain insight into JYNNEOS’ immunogenicity and mpox protection, we evaluated various metrics of adaptive responses in US adults after one and two JYNNEOS vaccine doses. These adults were all born after 1974 and were not exposed to smallpox or smallpox vaccines (as the US ended routine vaccinations in 1972). A strength of this study is that we used paired samples to evaluate B cell responses at the single cell and serum levels, combined with cellular antigen-specific T cell analyses.

Several lines of evidence in this study suggest that JYNNEOS protection is not primarily mediated by B cells in humans: the diversity, mutation levels, and CDR3 lengths of B cell heavy chain sequences revealed no significant changes after two doses. Vaccines mediated by antibodies usually induce changes in antibody genes that increase their binding and functional activity^43,53–57^. VH gene usage also usually changes after human immunization^58–60^ but did not significantly increase or decrease in the six gene families we analyzed.

Additionally, gene expression data revealed no significant changes after vaccination, although only analyzed in one individual at each time point (same subject for pre-vaccination and post-dose 1). A recent study mapping the expression patterns of single plasmablasts from > 800 adults in response to 13 vaccines^61^ confirmed that there is no single universal marker of vaccination in human plasmablasts. Our dimensionality reduction analyses by UMAP revealed that genes related to B cell activation, such as CD74, were increased in plasmablasts after one dose compared to in B cells in the pre-vaccination samples, as expected. However, consistent with our BCR sequencing results, we did not observe any gene expression changes between post-dose one and two, confirming that JYNNEOS did not significantly impact human B cell responses.

OPXV-elicited antibodies have significantly better cross-neutralizing potential compared to those targeting other viruses, such as HIV and influenza virus^26^, due to the a) high sequence homology between the surface proteins of VACV, mpox virus, smallpox virus, and cowpox virus (89–100% similarity) and b) the broad neutralization effects of OPXV antibodies, which can target multiple viral surface proteins concomitantly^62–64^. Therefore, we evaluated antibody secretion by memory B cells, with the idea that if serum antibodies offered direct protection against the virus in circulation, we could combine this knowledge with the single-cell data to better characterize humoral responses to JYNNEOS in humans.

However, serum IgG antibodies in post-vaccination samples did not bind any of the recombinant mpox proteins. Since cross-reactive antibodies are expected in response to OPXV exposure^38^, we tested the same sera against vaccinia proteins. Again, neither one nor two doses of JYNNEOS produced antibodies that could bind the vaccinia proteins, suggesting that the JYNNEOS vaccine cannot elicit strong humoral responses post-vaccination. The strong IgG responses to the same OPXV proteins by sera from subjects 20–102 days post-mpox infection clearly illustrate the vaccine’s limited antibody response. Convalescent participants presented a range of IgG responses against the proteins we analyzed, consistent with studies showing that different viral loads and clinical determinants dictate the response to mpox virus infection^65^. A recent ELISA-based study reported vaccine-specific IgG peaks in the serum of JYNNEOS-vaccinated participants after 42 days^66^. In addition, a recent study reported lower A35R- and H3L-specific titers in antigen-specific IgG tests compared to tests against the whole sera of convalescent participants, suggesting that IgMs might be among the immunoglobulins responding up to 50 days post-viral exposure (in that case, *via* infection)^40^. The reference strains of VACV and mpox virus share > 90% identity. The 2022 mpox virus strain has unique mutations compared to the reference strain, including A35R mutations at positions A67 (to L) and A88 (to V), which were exclusively found during the 2022 outbreak^38^. Whether these mutations can impact serum IgG binding to the recombinant mpox virus protein remains to be investigated. We found a positive correlation between JYNNEOS-elicited antibodies against the A35R and A33R proteins, suggesting that the common sequences these orthologs share induce comparable antibody responses to the vaccine. Finally, we confirmed the A35R protein as a serum marker of human mpox virus infection^40^, given the total absence of AS35R-IgG in the serum after vaccination.

Our data establish that the B cell immunogenicity of JYNNEOS vaccination is low up to 2 months post-vaccination.Thus, what mediates JYNNEOS’ protective effects in humans if not B cells and antibodies?

Immunity to many pathogens, including viruses, can be highly dependent on T cells^67,68^. To test if JYNNEOS induces T cell-dependent immune protection, we tested T cell responses against peptide pools designed based on VACV antigens, although in a limited set of samples due to lack of cell availability. Two doses of JYNNEOS elicited substantial CD8+, CD4+, and cTFH T cell responses with similar fold increases. We also noted the presence of pre-existing CD4+, but not CD8+, T cell immunity, as previously reported for Dryvax vaccination^30^. JYNNEOS vaccination and mpox infection produced similar magnitudes of reactivity for CD4+ and CD8+ T cells; however, after infection, the frequency of the T cell response trended lower after infection than after vaccination, consistent with a previous study showing that mpox infection specifically decreased antiviral-specific T cell reactivity^69^. Intriguingly, we also observed significantly lower cTFH T cell induction after infection versus vaccination. Finally, the quality of the vaccinia-specific T cells indicates the presence of a Th1/Th2 mixed phenotype, as previously reported for smallpox vaccination^70^ but not, until now, in the context of mpox virus infection. Our study was not designed to address particularities across different vaccinia virus-based vaccination platforms. Accordingly, whether the reduced antibody activity seen in response to JYNNEOS is related to inferior neutralization activity compared to Dryvax^71^ needs further assessment. Additionally, although previous reports demonstrated that the antigens recognized by T cells are highly correlated with those recognized by antibody responses^42^, JYNNEOS’ T cell-mediated protection might be due to epitopes elicited only in response to vaccinia-based vaccines and not mpox infection as the two proteomes share 61-67% of sequence conservation^30^. Alternatively, genes lost in the MVA versus Dryvax^72^ may account for a fraction of T cell responses that are not induced in JYNNEOS vaccination. Future studies should address, more in-depth, the antigens recognized by T cell responses.

This study has some limitations. We did not assess the roles of other immune cells (*e*.*g*., innate immune cells) in mediating mpox protection. The gene expression data was assessed using one subject per time point; however, in our opinion, this does not reduce the importance of the data, which was used for descriptive purposes, with no statistical comparisons performed between the different groups. Our study did not comprise virus neutralization assays, and thus, we cannot assess whether the low humoral response would be associated with low viral neutralization.^713072^

Taken together, our results indicate that JYNNEOS elicits low B cell and antibody responses in humans. However, it induces a robust T cell response that can recognize mpox virus and VACV peptides. These data provide insights into the protective mechanisms of a third-generation OPXV vaccine, which can be used to inform vaccine design and clinical data assessment during future OPXV outbreaks.

## Data Availability

All data produced in the present work are contained in the manuscript

## Author contributions

CHC and VS conceptualized the study. HC, NB, JC, and AT performed the experiments. JC, WR, and LQ designed, expressed, and isolated recombinant mpox proteins. NB, JC, HC, AT, and CHC analyzed the data. FPL and SM performed bioinformatic analyses. CHC, SC, and VS funded the study. CHC, VS, SC, AG, AS, and FK supervised experiments. VS supervised the clinical study. CHC wrote the manuscript draft. All authors interpreted the data, provided critical input, and revised the manuscript.

## Data Sharing

Additional deidentified data may be made available to investigators whose proposed use of the data has been approved by the Icahn School of Medicine Ethics Review Board upon request to the corresponding authors.

## Acknowledgments

Research reported in this publication was supported by the National Cancer Institute of the National Institutes of Health under Award Number U54CA267776. We thank the Sequencing Core Facility at the La Jolla Institute for Immunology for sequencing the cDNA libraries. The NovaSeq 6000 was acquired through the Shared Instrumentation Grant Program (S10; 6000 S10OD025052). Substantive, stylistic, and copy editing of the manuscript and supplemental materials were provided by Dr. Nicole St. Denis at High-Fidelity Science Communications (www.hifiscicomm.ca). This project has been funded in whole or in part with Federal funds from the National Institute of Allergy and Infectious Diseases, National Institutes of Health, Department of Health and Human Services, under Contract No. 75N93021C00065 to A.S.

