## Supplementary Material for "Mpox vaccine and infection-driven human immune signatures"

### Supplementary Figures

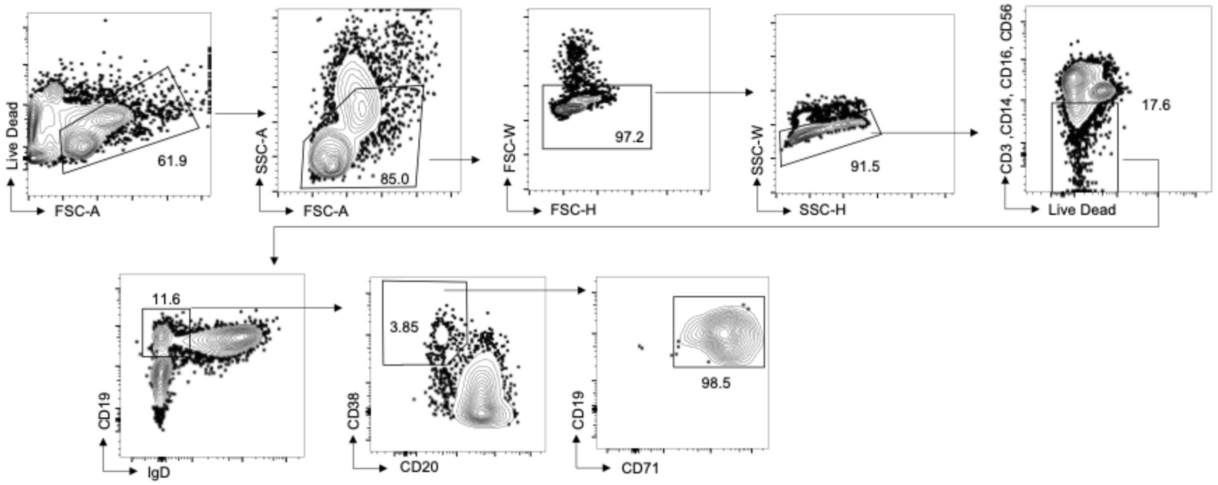


**Figure S1. Flow cytometry gating strategy to identify and sort plasmablasts from the PBMCs of JYNNEOS recipients**

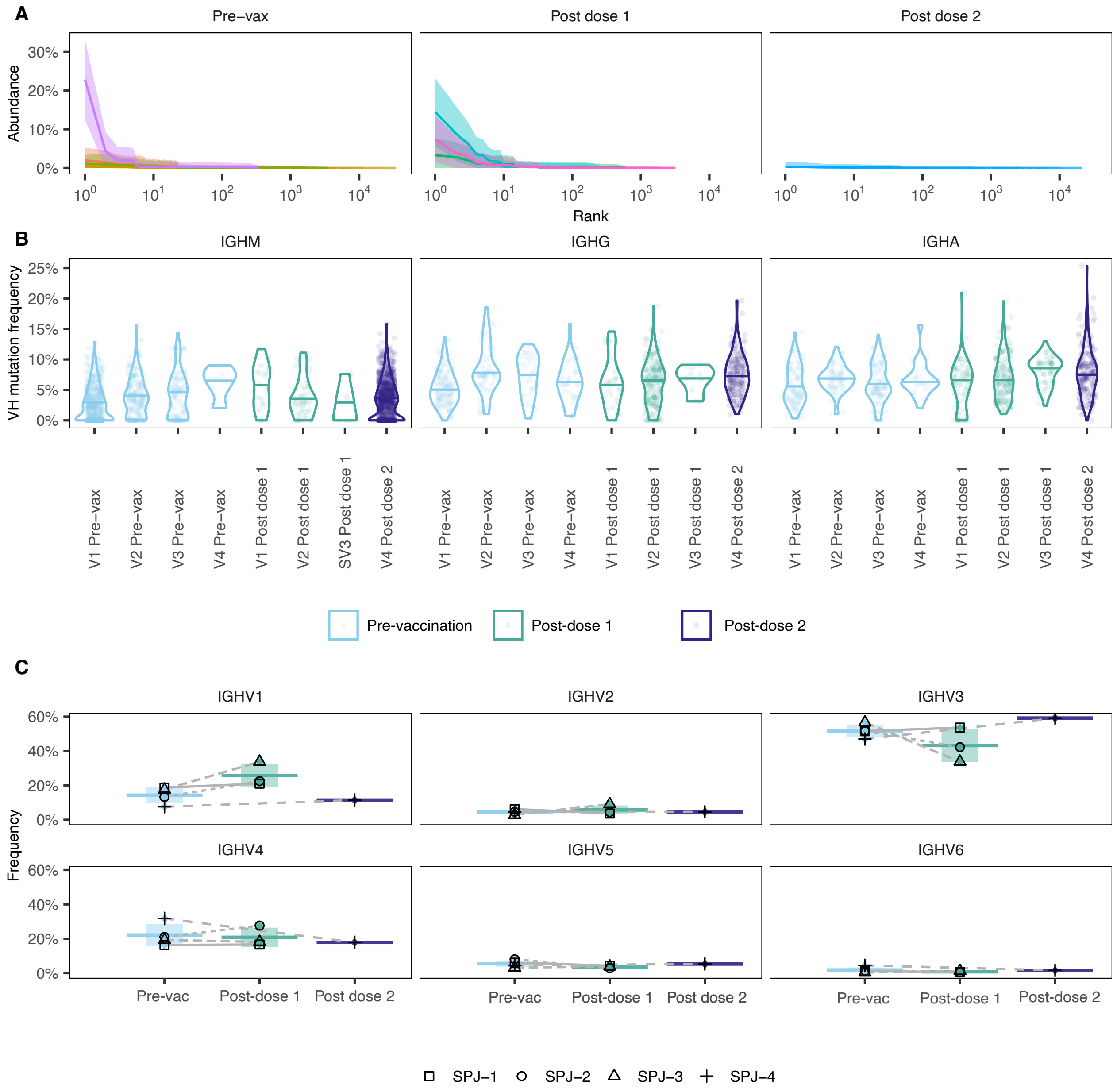


**Figure S2. Antibody repertoire induced by JYNNEOS vaccination**

**(A)** Abundance of heavy chain clonotypes in plasmablasts pre-vaccination and after 1 and 2 doses.

**(B)** Mutations by immunoglobulin isotype (visualized by individual).

**(C)** VH gene family usage.

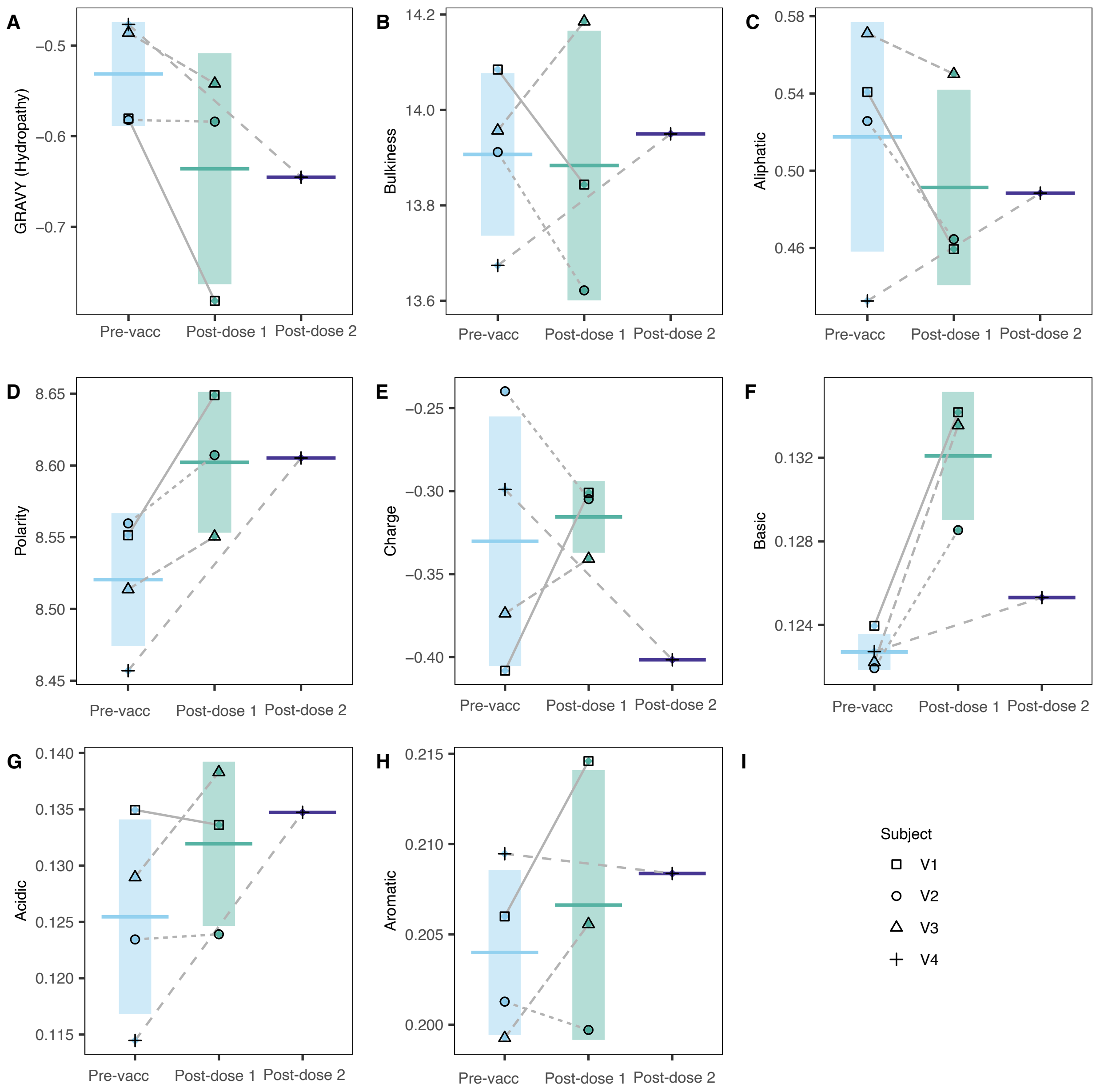


**Figure S3.** Physicochemical properties of complementarity-determining region 3 in human B cells pre-vaccination and after one and two doses of JYNNEOS.

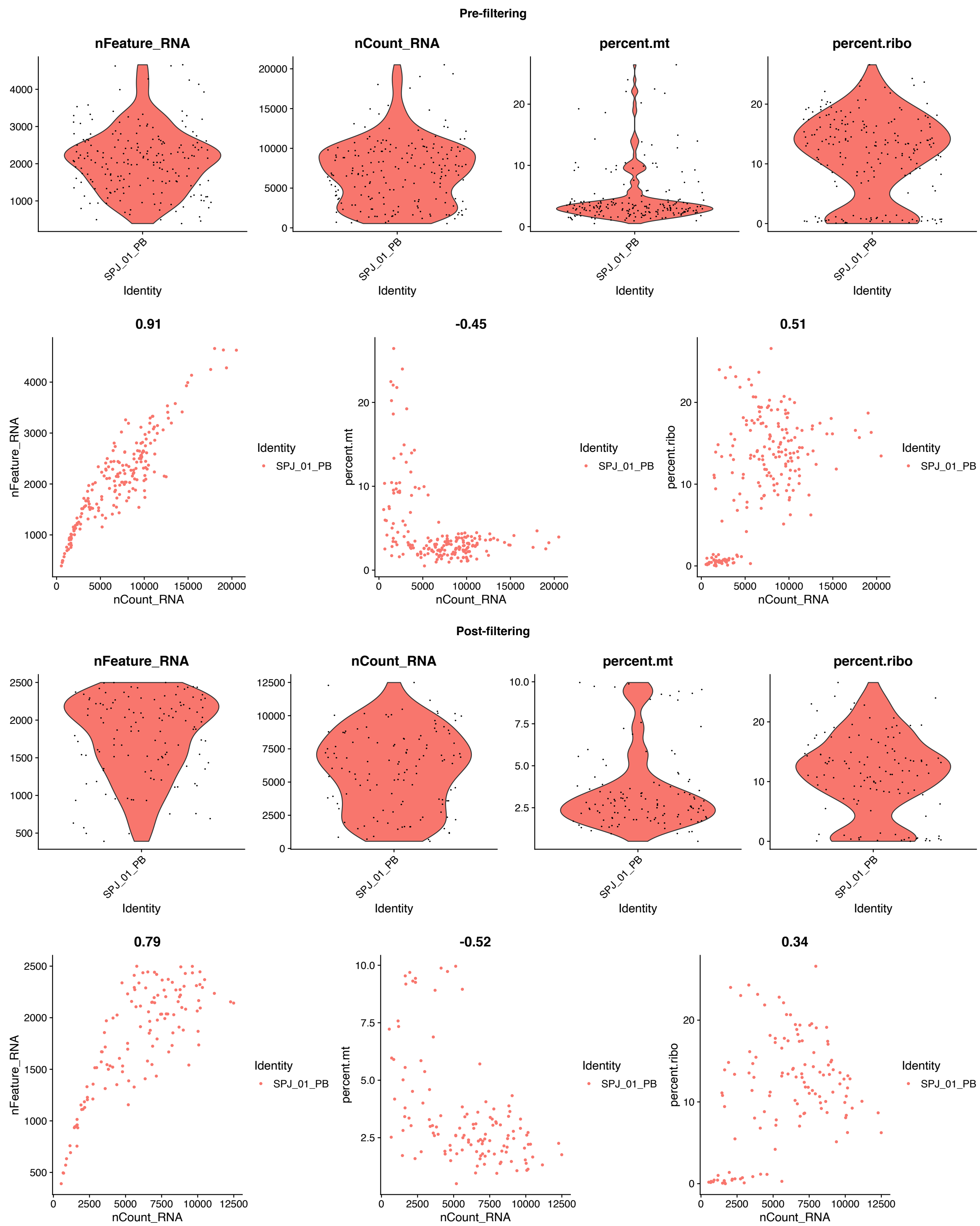


**Figure S4. Quality control of single-cell RNA sequencing data for gene expression analyses in plasmablasts. .** Individual cell barcodes are represented as red dots. Data types for individual cell barcodes are as follows: nFeature_RNA - number of transcribed loci; nCount_RNA: total number of reads mapping in transcribed loci; percent.mt - percentage of reads mapping to mitochondrial loci; percent.ribo - percentage of reads mapping to ribosomal protein genes. We used the following cutoffs to select high-quality cells: percent.mt < 10%; 200 < nFeature_RNA < 2500. Data for subject V1 are shown. The analyses were performed on all samples.

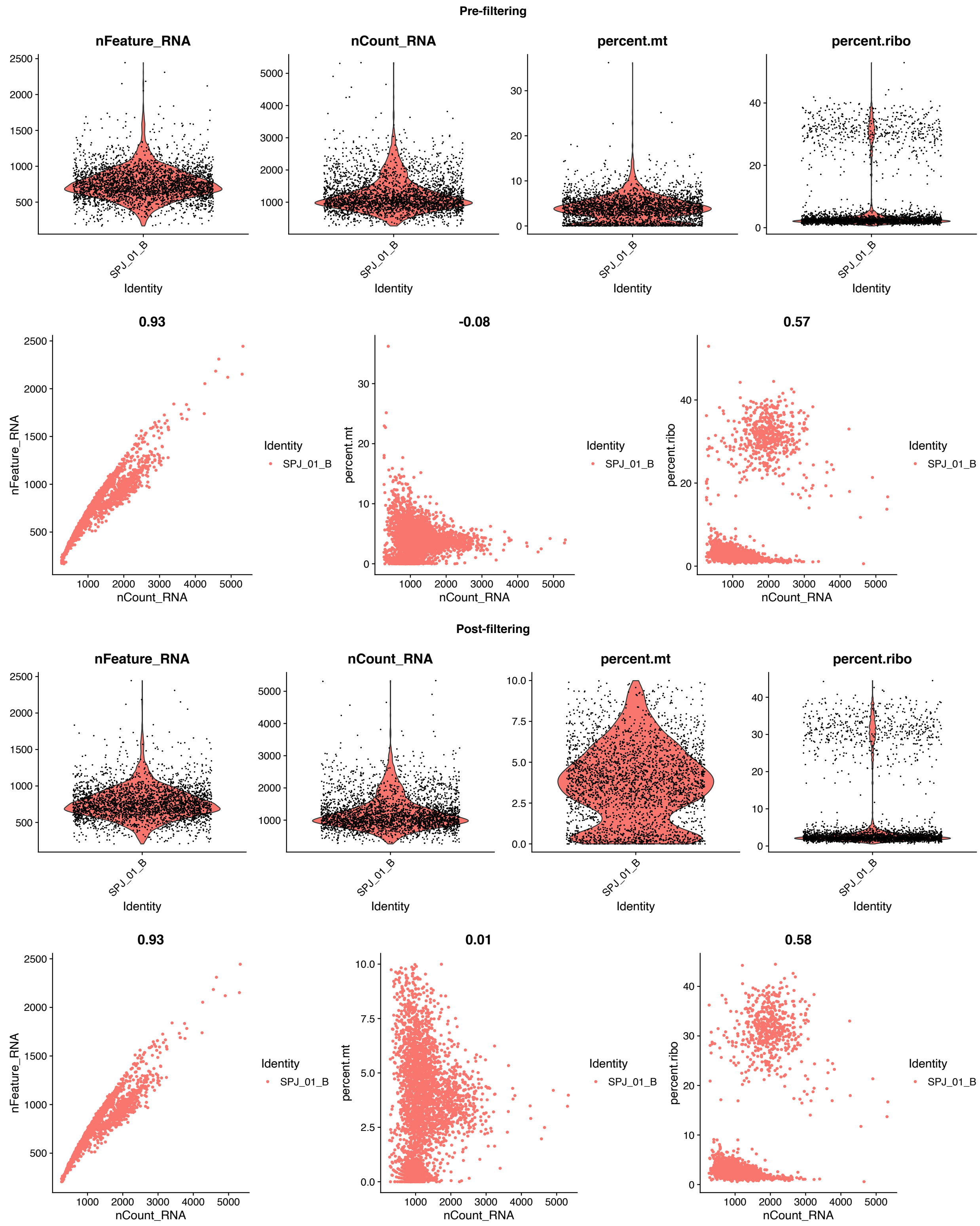

**Figure S5. Quality control of single-cell RNA sequencing data for gene expression analyses in total B cells (CD19+)** Data for subject V1 are shown. The analyses were performed on all samples. See Figure S4 for the definition of data types.

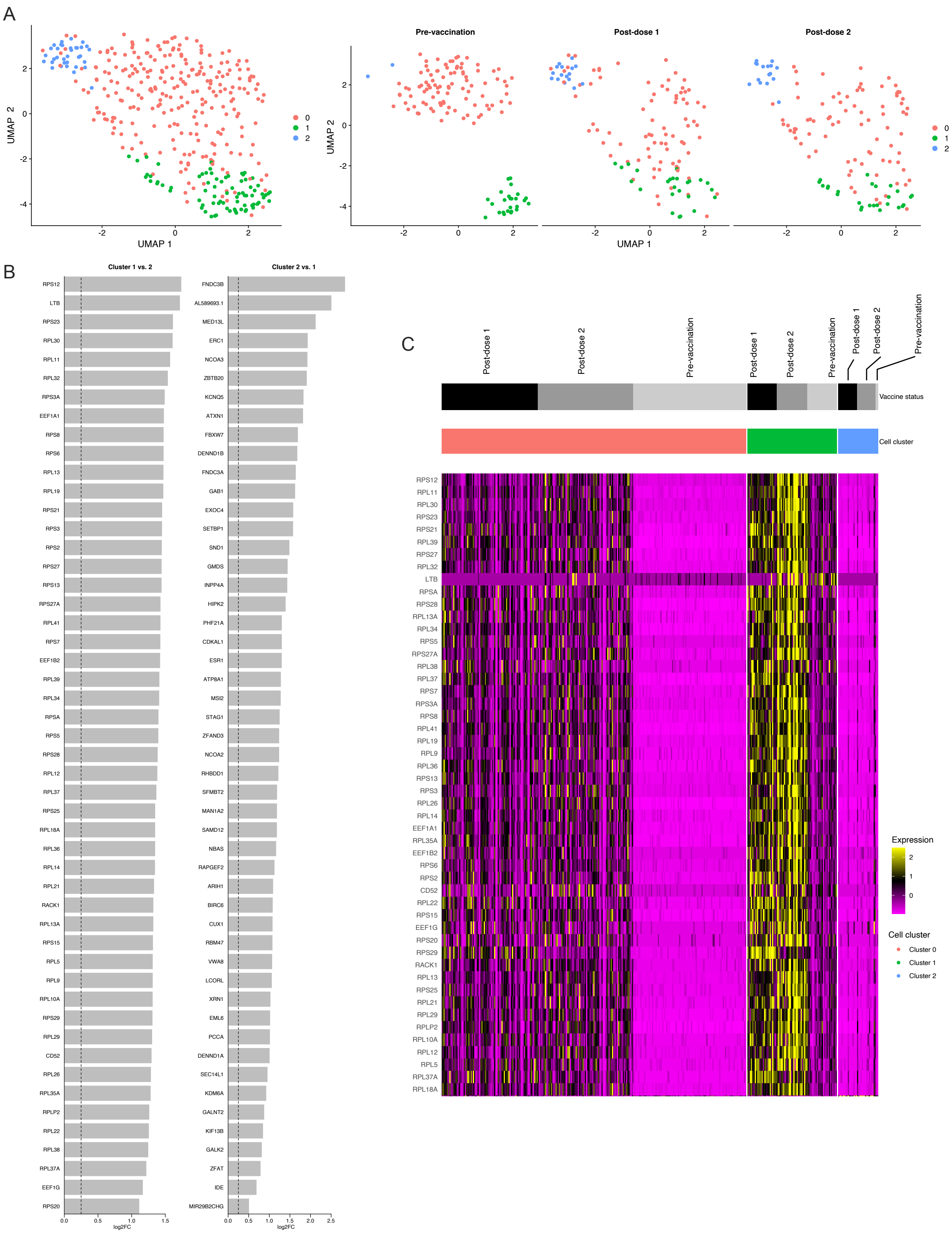
A
UMAP 1
UMAP 2
0
1
2
Pre-vaccination
Post-dose 1
Post-dose 2
B
Cluster 1 vs. 2
RPS12
LTB
RPS23
RPL30
RPL11
RPL32
RPS3A
EEF1A1
RPS8
RPS6
RPL13
RPL19
RPS21
RPS3
RPS2
RPS27
RPS13
RPS27A
RPL41
RPS7
EEF1B2
RPL39
RPL34
RPSA
RPS5
RPS28
RPL12
RPL37
RPS25
RPL18A
RPL36
RPL14
RPL21
RACK1
RPL13A
RPS15
RPL5
RPL9
RPL10A
RPS29
RPL29
CD52
RPL26
RPL35A
RPLP2
RPL22
RPL38
RPL37A
EEF1G
RPS20
log2FC
Cluster 2 vs. 1
FNDC3B
AL589693.1
MED13L
ERC1
NCOA3
ZBTB20
KCNQ5
ATXN1
FBXW7
DENND1B
FNDC3A
GAB1
EXOC4
SETBP1
SND1
GMDS
INPP4A
HIPK2
PHF21A
CDKAL1
ESR1
ATP8A1
MSI2
STAG1
ZFAND3
NCOA2
RHBDD1
SFMBT2
MAN1A2
SAMD12
NBAS
RAPGEF2
ARIH1
BIRC6
CUX1
RBM47
VWA8
LCORL
XRN1
EML6
PCCA
DENND1A
SEC14L1
KDM6A
GALNT2
KIF13B
GALK2
ZFAT
IDE
MIR29B2CHG
log2FC
C
Post-dose 1
Post-dose 2
Pre-vaccination
Post-dose 1
Post-dose 2
Pre-vaccination
Post-dose 1
Post-dose 2
Pre-vaccination
Vaccine status
Cell cluster
RPS12
RPL11
RPL30
RPS23
RPS21
RPL39
RPS27
RPL32
LTB
RPSA
RPS28
RPL13A
RPL34
RPS5
RPS27A
RPL38
RPL37
RPS7
RPS3A
RPS8
RPL41
RPL19
RPL9
RPL36
RPS13
RPS3
RPL26
RPL14
EEF1A1
RPL35A
EEF1B2
RPS6
RPS2
CD52
RPL22
RPS15
EEF1G
RPS20
RPS29
RACK1
RPL13
RPS25
RPL21
RPL29
RPLP2
RPL10A
RPL12
RPL5
RPL37A
RPL18A
Expression
Cell cluster
Cluster 0
Cluster 1
Cluster 2

**Figure S6. Gene expression before (CD19+ cells) and after one or two doses of JYNNEOS (plasmablasts)**
**(A)** UMAP analyses showing the two clusters identified before and after vaccination.
**(B)** The top differentially expressed genes in clusters 0 and 1 are shown.
**(C)** Heatmap of the expression intensities of the differentially expressed genes in each cluster.

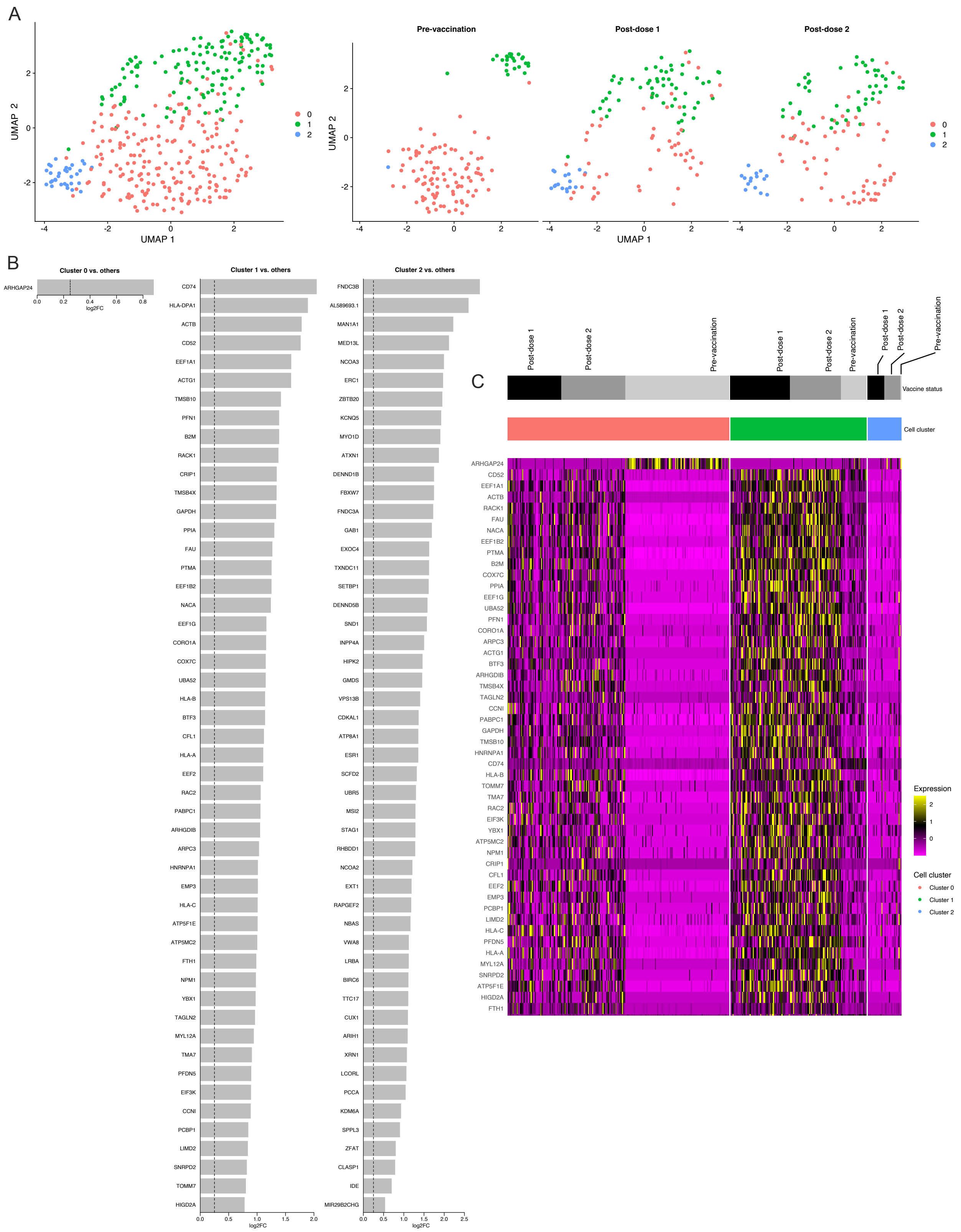

**Figure S7. Gene expression before (CD19+ cells) and after one or two doses of JYNNEOS (plasmablasts), with ribosomal proteins removed**

**(A)** UMAP analyses showing the two clusters identified before and after vaccination

**(B)** The top differentially expressed genes in clusters 0 and 1 are shown. . We removed the counts of reads mapping for genes coding ribosomal proteins from the Seurat objects containing the gene expression raw count matrix gathered from the Cell Ranger output. Genes coding for ribosomal proteins were defined as those with gene symbols beginning with "RP" (some examples can be seen in Figures S6B-C).
**(C)** Heatmap of the expression intensities of the differentially expressed genes in each cluster.

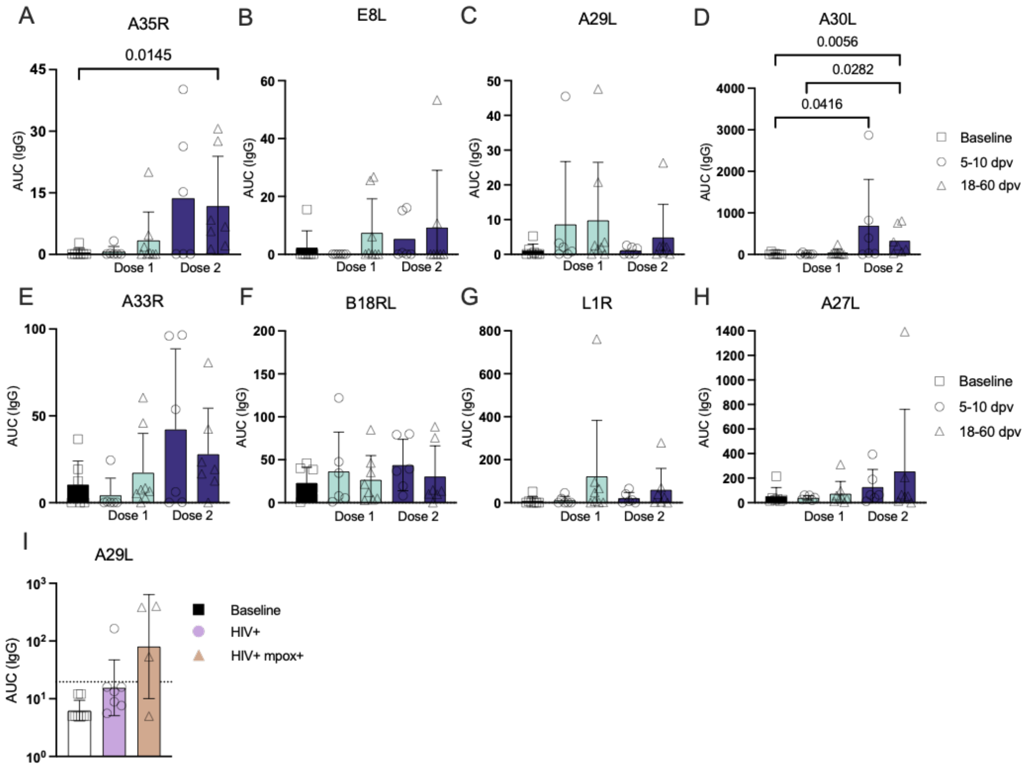


**Figure S8.** Serum antibody responses to mpox **(A-D)** and vaccinia **(E-H)** recombinant proteins after JYNNEOS immunization (visualized 5–10 and 18–60 days post-vaccination (dpv)) or mpox infection. **(I)** HIV+ and HIV+/mpox+ serum antibody responses to the mpox recombinant protein A29L.The limit of detection equals 19.58597 and was calculated using the mean of the baseline + 3 standard deviations. Comparisons were performed using the Kruskal-Wallis test followed by Dunn's multiple comparisons test (**p* < 0.05, ***p* <0.01, ****p* < 0.001).

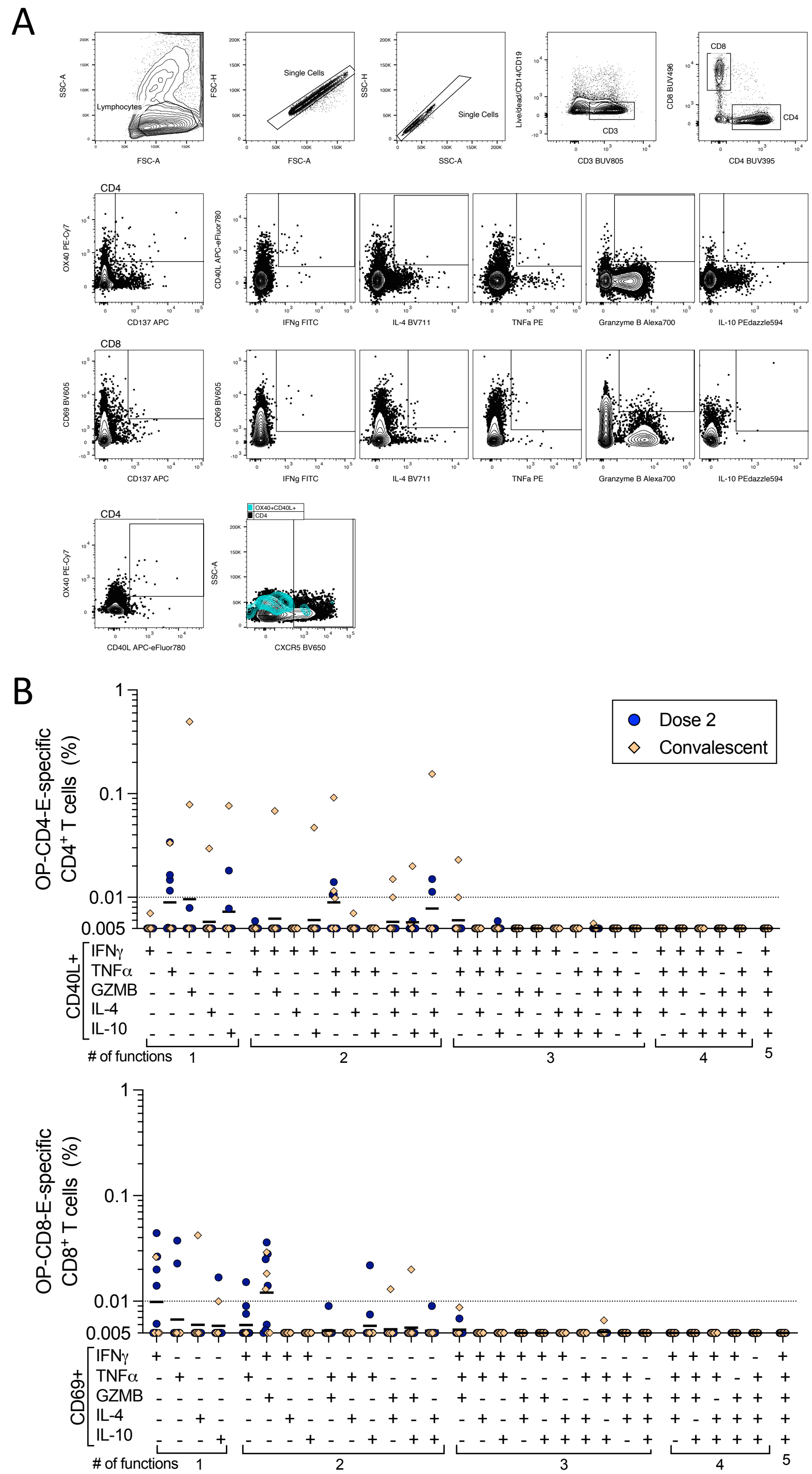


**Figure S9.** T cell assay gating strategy and orthopox-specific T cell cytokine compositions

**(A)** AIM/ICS flow cytometry assay gating strategy to identify orthopox-specific T cells.

**(B)** Boolean analysis of IFNγ, TNFα, granzyme B, IL4, and IL10 production by orthopox-specific CD4 and CD8 T cells after JYNNEOS vaccination or mpox infection. The dotted line indicates the limit of sensitivity.

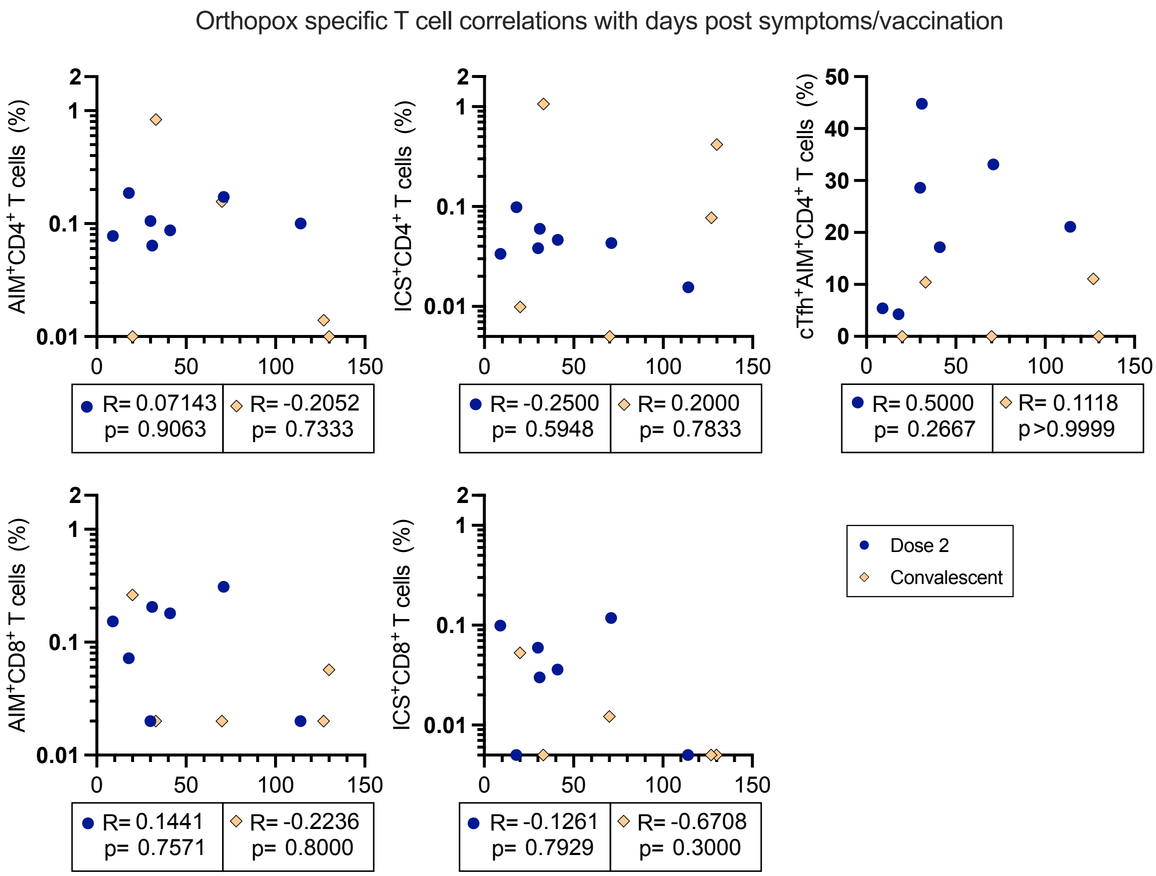


**Figure S10. Correlations between T cell responses and the time post-vaccination or post-infection**

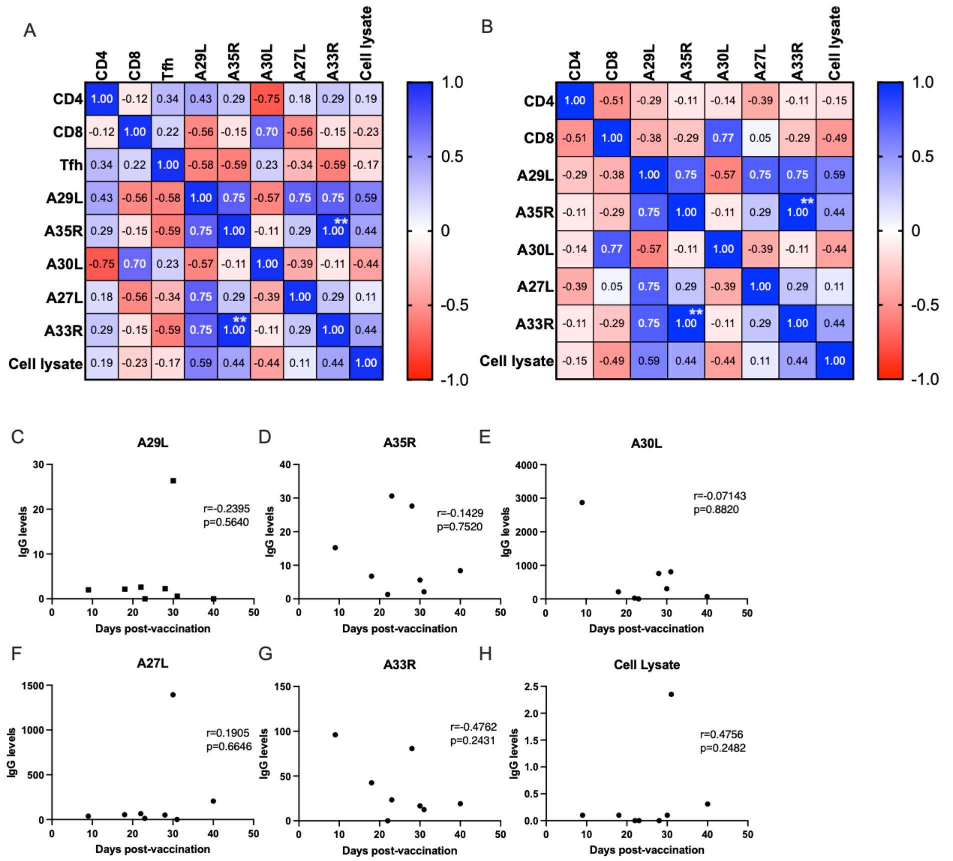


**Figure S11. Correlations between T cells, antibody titers, and the time post-vaccination**
**(A-B)** Correlations between the CD4, CD8, and Tfh T cell responses and the antibody responses after two doses of JYNNEOS
**(C–H)** Spearman's correlations between the antibody titers of orthopox proteins and the days post-vaccination. Analyses were performed with all viral proteins. Only proteins leading to serum IgG responses are shown

### Supplementary Tables

**Table S1. Number of variable heavy chain (VH) sequences recovered after sorting total B cells and plasmablasts.**

| Subject ID | Sample Type | Days post-immunization | Cell type | Number of cells sorted | Number of VH sequences recovered |
|---|---|---|---|---|---|
| **V1** | pre-vaccination | 0 | Total B cells | 10,000 | 850 |
| **V1** | post-dose 1 | 6 | Plasmablasts | 569 | 138 |
| **V2** | pre-vaccination | 0 | Total B cells | 10,000 | 370 |
| **V2** | post-dose 1 | 7 | Plasmablasts | 705 | 459 |
| **V3** | pre-vaccination | 0 | Total B cells | 5,662 | 279 |
| **V3** | post-dose 1 | 7 | Plasmablasts | 537 | 77 |
| **V4** | pre-vaccination | 0 | Total B cells | 10,000 | 66 |
| **V4** | post-dose 2 | 9 | Plasmablasts | 1,307 | 1,532 |

**Table S2. Viability (in %) of human PBMCs used in combined AIMS/ICS assays.**

| Sample ID | Pre-vaccination | Post-dose 2 | Convalescent |
|---|---|---|---|
| **V1** | 82% | 85% | - |
| **V2** | 94% | 93% | - |
| **V4** | 94% | 83% | - |
| **V5** | 69% | 70% | - |
| **V6** | 83% | 95% | - |
| **V8** | 87% | 82% | - |
| **V10** | 87% | 90 | - |
| **V11** | 94% | 90% | - |
| **C2** | - | - | 75% |
| **C3** | - | - | 96% |
| **C4** | - | - | 83% |
| **C8** | - | - | 83% |
| **C9** | - | - | 81% |
